# Undiagnosed Hypertension and Its Associated Factors: A Cross-Sectional Study among Adults in a Rural Municipality of Nepal

**DOI:** 10.1101/2025.11.25.25340960

**Authors:** Anil Shrestha, Bipsana Shrestha, Bhoj Raj Kalauni, Ram Hari Regmi, Prabin Karki, Kamal Ranabhat, Buna Bhandari

**Affiliations:** Central Department of Public Health, Institute of Medicine, Tribhuvan University, Kathmandu, Nepal; Health Directorate, Koshi Province, Dhankuta, Nepal; Department of Health Services, Ministry of Health and Population, Kathmandu, Nepal; Ministry of Health and Population, Kathmandu, Nepal; Florida State University College of Nursing, Tallahassee, Florida, USA; Department of Global Health and Population, Harvard TH Chan School of Public Health, Boston, USA

**Author notes:** Anil Shrestha (AS). Bipsana Shrestha (BS). Ram Hari Regmi (RHR). Prabin Karki (PK). Kamal Ranabhat (KR). Buna Bhandari (BB). **Corresponding author:** Anil Shrestha, Central Department of Public Health, Institute of Medicine, Tribhuvan University, Nepal.

**Keywords:** high blood pressure, screening, treatment, prevention

## Abstract

**Introduction:** Hypertension is a silent killer. Early detection of hypertension is crucial for effective management. Yet, the condition remains unnoticed until complications emerge, especially in resource limited setting.

**Objective:** The main objective of this study was to assess prevalence of undiagnosed hypertension and its associated factors among adults residing in Jwalamukhi Rural Municipality, Dhading, Bagmati Province.

**Methods:** A cross-sectional study was conducted among the individuals aged 18 years and above between November 10 and May 10, 2024, in a Rural Municipality of Dhading district, Nepal. A multistage sampling method was used to select 422 study participants. Data were collected using a structured, STEPwise questionnaire, entered into EpiData 3.1, and analyzed using IBM SPSS version 20. Descriptive analysis and Chi-square or Fisher’s exact tests was used to assess the factors associated with undiagnosed hypertension a. Predictors with p<0.05 were chosen for further multivariate analysis. Odds ratio was utilized to evaluate the strength of relationships (95% CI), where a p-value of <0.05 indicated statistical significance.

**Results:** The Prevalence of undiagnosed hypertension was found to be 30.80% [95% CI: 26.4-35.5] where Male [AOR=2.2, 95% CI: 1.1-4.2], Salt consumption of more than 5 grams a day [AOR=3.1, 95% CI: 1.8-5.4], and blood pressure not measured regularly [AOR=5.5, 95% CI: 3.1-9.8] was associated with undiagnosed hypertension.

**Conclusion:** This study revealed a high prevalence of undiagnosed hypertension, underscoring the urgent need for comprehensive and effective screening strategies to detect and manage hypertension in the broader population of Nepal.

## Introduction

Hypertension (HTN) is a major modifiable risk factor for many Non-Communicable diseases, such as cardiovascular disease, diabetes, and chronic renal disease ^1^. Approximately 1.4 billion individuals between the ages of 30 and 79 worldwide are impacted by hypertension, with the majority, around two-thirds, residing in low- and middle-income nations ^2^. It is expected to reach 1.5 billion by 2025 ^3^.

Globally, Hypertension is accountable for about 7.5 million fatalities, or nearly 12.8% of the total mortality. It also leads to 57 million disability-adjusted life years (DALYs), accounting 1.5 million death for 3.7% of the total DALYs ^4^. Overall, Non-communicable diseases (NCDs), including hypertension, caused 41 million individual deaths globally, making up 74% of all fatalities ^5^.

One of the goals set by WHO regarding non-communicable illness is to reduce the prevalence of hypertension by 33% between 2010 and 2030 ^6^. Similarly, the Sustainable Development Goal (SDG) 3.4 seek to decrease premature deaths from NCDs by one-third via prevention and treatment as well as enhance mental health and wellbeing ^7^.

Importantly, approximately 46% people have undetected hypertension globally ^8^. Undiagnosed hypertension poses a significant public health concern in both in high income nations and lower to middle incomes countries (LMICs). This is due to various factors such as lifestyle choices, economic circumstances, and insufficient awareness across different socioeconomic backgrounds ^9^. Similarly, one in three adults in Southeast Asian countries is also affected by hypertension and it causes about 1.5 million fatality annually, which is 9.4% of all deaths, while more than 50% of hypertensive patients go undetected ^10^. In South-East Asia, over half of the population affected by hypertension remains undiagnosed ^11^. Where, Nepal exhibits one of the highest hypertension prevalence rate ^12^. Nepal at present, is experiencing an epidemiological transition with the increasing cases of undiagnosed hypertension ^13^.

The NDHS 2022 report indicated that the prevalence of hypertension varied between genders, with rates of 23 % for males and 18 % for females. Moreover, it revealed that 48% women and 52% men who were 15 and older in age, with hypertension were unaware of their condition ^14^. Another national representative survey conducted in 2019 showed that 24.5% of individuals aged 15–69 years had hypertension in Nepal. This percentage increased from 23.4% in 2013 to 24.5% in 2019, with 78.8% of adults being unaware of their elevated blood pressure ^15^. Although national survey reports such as the NDHS 2022 (NDHS 2002 ref) and the WHO STEPS Survey 2019 have shown that a large proportion of Nepalese adults remain unaware of their high blood pressure, very little is known about the situation in local communities. In rural settings such as Jwalamukhi Rural Municipality of Dhading district, differences in lifestyle, health-service reach, and living environment may lead to patterns of undiagnosed hypertension that differ from national trends. The survey also identified various factors for hypertension in Nepal, such as low physical activity, consumption of unhealthy food, use of tobacco, and alcohol use ^15^. Research on undiagnosed hypertension in Nepal is limited. However, existing studies have identified that undiagnosed hypertension is more common among men, individuals living in Sudurpashchim Province, those from lower socio-economic groups, and people with variations in body mass index. Delayed diagnosis of hypertension can lead to serious yet preventable outcomes, including stroke, heart attack, and kidney. In Nepal, studies show that more than half of people living with high blood pressure are unaware of their condition, highlighting the urgent need for regular and systematic screening to support early detection and appropriate management. The change in illness patterns in Nepal may be ascribed to numerous causes, such as increased urbanization, changes in food habits, behavioral variables, and major advances in maternal and child health prevention, which have increased life expectancy ^16^.

Blood pressure screening can help detect this condition early and decrease the chance of additional complications ^17^. Hypertension is often called the “silent killer” because most people who have it experience no noticeable symptoms until serious complications develop. Evidence from large international studies suggests that this silent progression is a major reason why many cases of high blood pressure remain undiagnosed across populations^18^. There are numerous studies on the occurrence of hypertension and its associated factors but very few have investigated undiagnosed in particular study sites in Nepal. Hypertension is a major public health concern and one of the leading causes of preventable heart disease, stroke, and kidney failure worldwide. A large number of cases remain undiagnosed, particularly in low- and middle-income countries such as Nepal, where regular screening and health-care access are limited. Although national surveys have reported the overall prevalence of hypertension, they provide little insight into the true extent of undiagnosed cases and the factors influencing them at the community level. This gap is even more pronounced in rural areas, where geographic and economic barriers often delay detection and treatment. Generating local evidence is therefore essential to design practical screening and prevention strategies that fit the realities of rural Nepal. With this aim, the present study assesses the prevalence of undiagnosed hypertension and its associated factors in the Dhading District of Bagmati Province, Nepal.

## Methods and Materials

### Study design, site

A community-based cross-sectional study was conducted in Jwalamukhi Rural Municipality, Dhading district of Nepal, from November 10 to May 10, 2024. It is one of the municipalities of Bagmati province in Nepal, and 100 km away from Kathmandu, the capital of Nepal. The total population of Jwalamukhi Rural Municipality was 21,338, of which 9,981 were male and 11,356 were female, including 7 wards and 5730 households in Jwalamukhi RM.

### Study population

The study population included adults aged 18 years and older who lived permanently (at least 6 months) in Jwalamukhi Rural Municipality and Pregnant, medically confirmed case of hypertension and currently receiving medication for hypertension and physically injured and serious mental health illness were excluded from the study.

### Sample Size and Sampling technique

The sample size was calculated by using Cochran’s formula, n = z^2^pq/d^2,^ with these assumptions (p=0.504, q= 0.496 and z= 1.96 at 95% confidence interval, and d= 0.05) ^11^. The total sample size of 422 individuals included 10% non-response rate.

A multistage random sampling method was used to choose four hundred twenty-two (n=422) participants. The study’s participants were recruited using a multistage sampling. Among 7 wards in the rural municipality, 3 wards were selected randomly (Ward 1, Ward 3, and Ward 4). Following this, the whole sample was divided among the chosen wards depending on the number of houses. Subsequently, houses were chosen from each ward using the probability proportional to size (PPS) method as a sampling framework. Then, through door-to-door surveys, one eligible member was recruited from each house. In cases where multiple eligible participants were present within a household, a single participant was selected through a lottery process.

**Fig 1:**
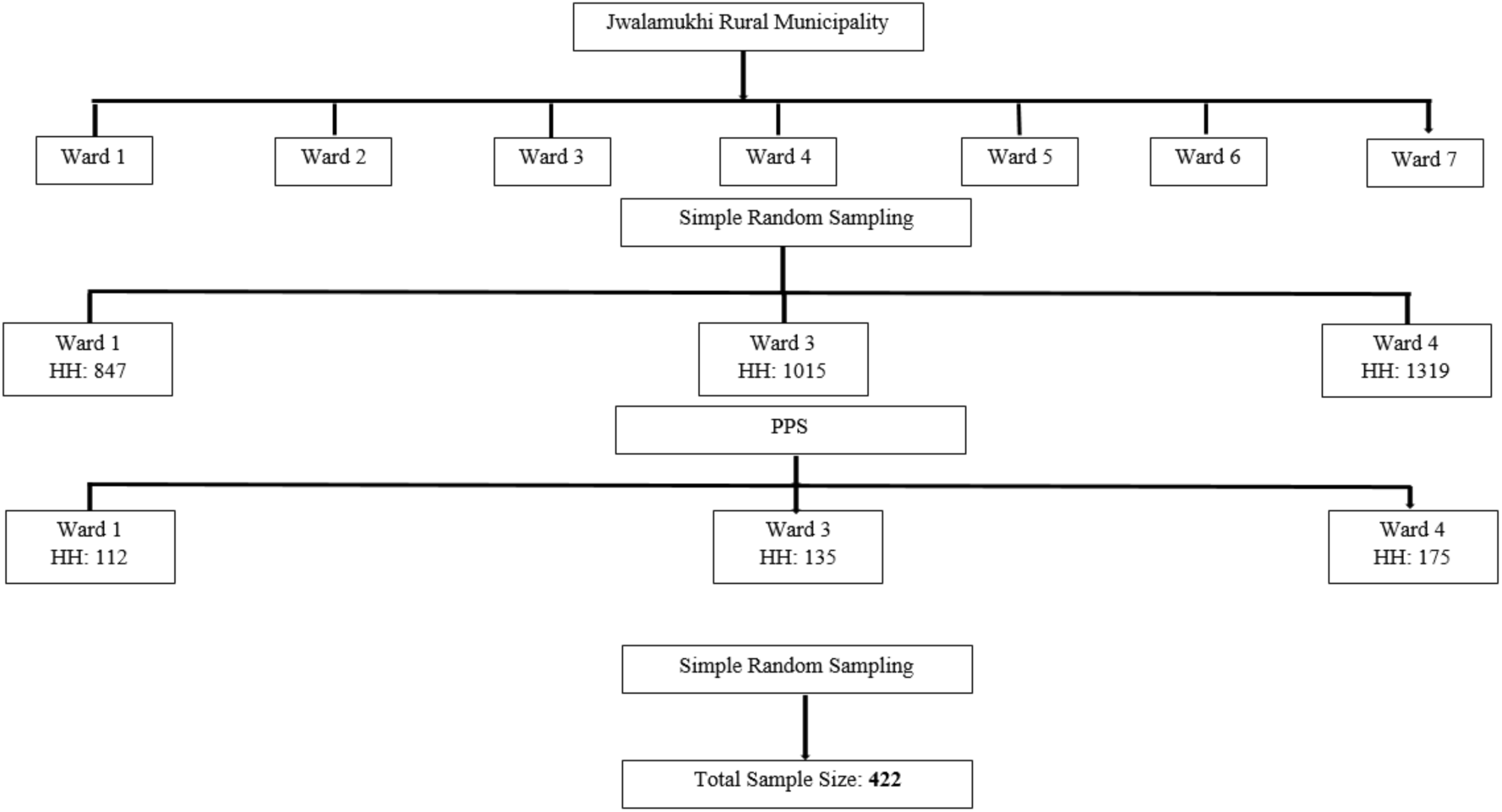
Schematic representation of the sampling technique.

### Study variables

The outcome variable of the study, undiagnosed hypertension, was split into either a yes or no group for binary logistic regression analysis. The independent variable includes sociodemographic factors, behavioral risk factors, clinical characteristics, healthcare-seeking behavior, and anthropometric measurements.

### Data collection instruments and techniques

The survey utilized the WHO-NCD Stepwise approach to surveillance tool to gather demographic characteristics, behavioral data, and anthropometric characteristics. In step 1: sociodemographic variables and behavioral risk factors were assessed and in step 2: Physical measurements (height, weight and BP) were taken.

### Physical measurements: Anthropometry and Blood pressure

#### Blood pressure

Blood pressure was measured using an OMRON digital monitor equipped with a universal-size cuff by principal investigator himself. Participants were requested to sit calmly with their legs uncrossed for 15 minutes before the measurements were taken. Three measurements of BP were obtained, with a 3-minute rest period between each reading. The mean of the 2^nd^ and 3^rd^ measurements was computed. The sphygmomanometer cuff was positioned on the participant’s left arm with resting forearm, palm facing upward…

#### Anthropometry

All sampled individuals underwent measurements for both height and weight. Height measurements were taken using a standard Stadiometer. Participants were instructed to remove any footwear, hats, or hair ties. They were then asked to stand barefoot on a plain surface, facing to interviewer with feet together and knees straight, maintaining an eye contact. Height readings were recorded in cm.

Weight was measured with SAMSO digital weighing scale. The scale was placed on a firm and flat terrain. Participants were requested for lightweight clothing. Then participants were requested to stand on the machine with one foot on each side, facing forward, with their arms at their sides. The weight was recorded in kg.

The body mass index (BMI) was calculated by dividing weight in kilogram by height in meters squared.

### Operational definitions Hypertension

Hypertension refers an increase BP ≥140/90 during a study. Participants who take anti-hypertensive drugs are also considered hypertensive ^19^.

### Undiagnosed Hypertension

Refers to blood pressure ≥140/90 during a survey, had not used any prescribed antihypertensive medication and never received a diagnosis of hypertension from healthcare professional prior to this study ^20^.

### A family history

Family history of hypertension is indicated if the participant’s first-degree relatives, such as parents, grandparents, or siblings, have been diagnosed with hypertension or are taking antihypertensive medication ^21^.

### BMI

BMI was calculated using measured heights and weights. The study participants were classified as underweight (BMI <18.5), normal (BMI between 18.5 to 22.9), overweight (BMI between 23.0 and 24.9), or obese (BMI ≥ 25) as per South Asian BMI Classification ^22^.

### Regular blood pressure measure

Regular blood pressure measurement involves assessing blood pressure at least once a year ^23^.

### Statistical analysis

Descriptive analysis was done by calculating frequency and percentage. Chi-square or Fisher exact tests examined the relationship between undiagnosed hypertension and independent variables. The p<0.05 were chosen for further multivariate analysis. The odds ratio was utilized to evaluate the strength of relationships (95% CI), where a p-value of <0.05 indicated statistical significance.

### Ethical consideration

Ethical clearance was obtained from the Institutional Review Committee (IRC) of the Institute of Medicine, Tribhuvan University (Ref: 311(6-11) E2). Participants were informed about the voluntary nature of their participation and were assured of their anonymity and the confidentiality of their information. Those identified as having hypertension were advised to consult a nearby health facility.

## Results

### Sociodemographic characteristics of study participant

The mean age of the study participant was 49.98 ± 17.03 years, while the majority of the study participants were under 60 years (69%), and more than half were female (67.3%) and less than half belonged to disadvantaged Janajati groups (41.2%). The majority were married (88.6%) and less than half had completed non-formal education (45.7%). Additionally, more than half of the participants were engaged in farming (58.3%).

**Table 1.**
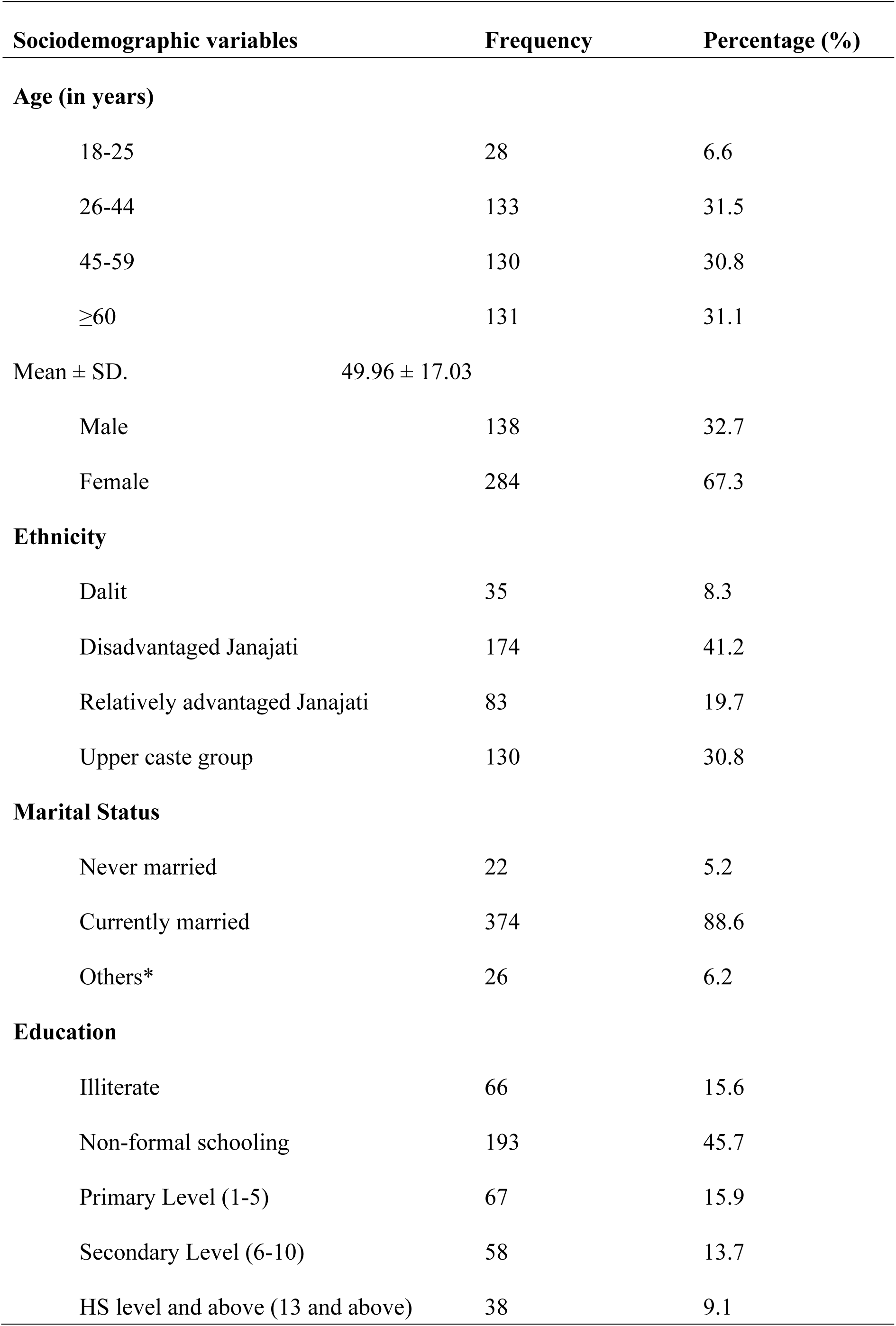
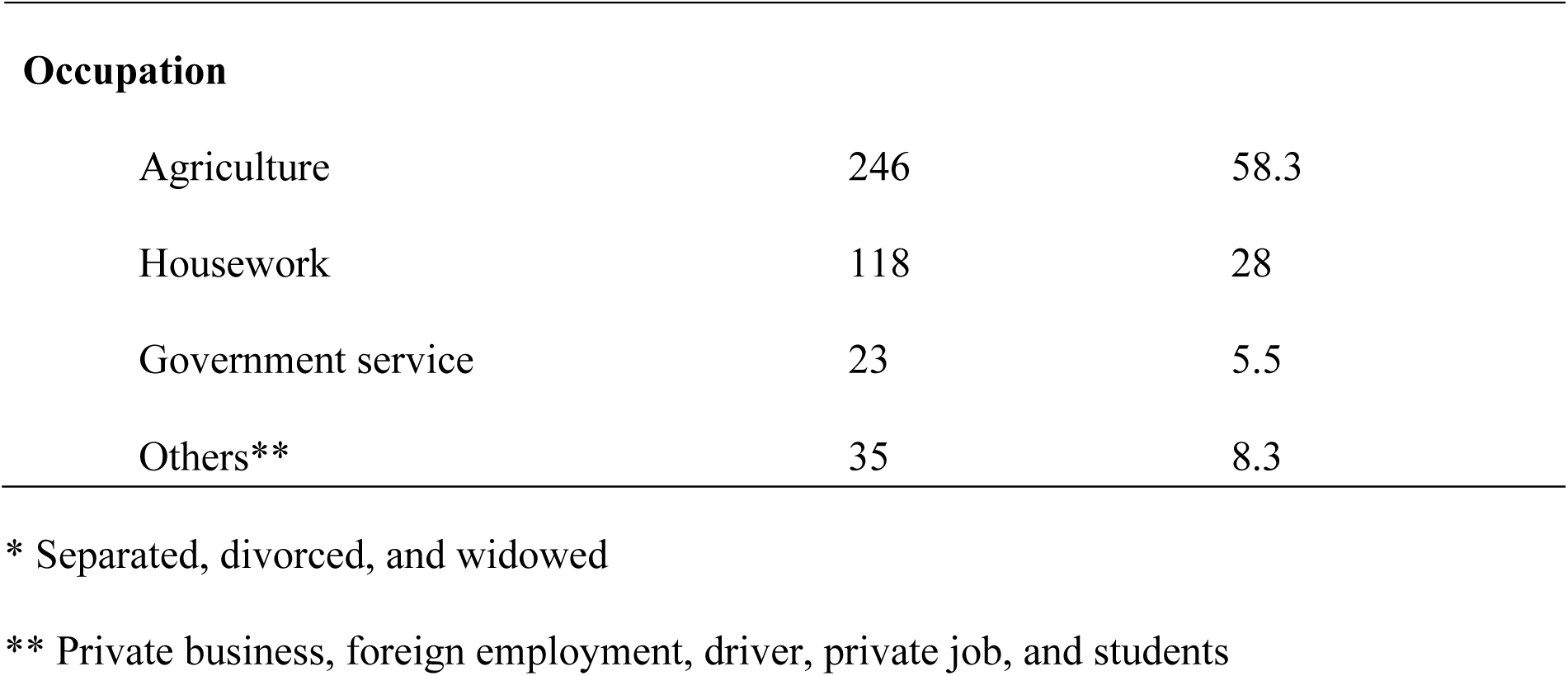
Sociodemographic characteristics of study participants (n=422)

| **Sociodemographic variables** | **Frequency** | **Percentage (%)** |
| --- | --- | --- |
| **Age (in years)** |  |  |
| 18-25 | 28 | 6.6 |
| 26-44 | 133 | 31.5 |
| 45-59 | 130 | 30.8 |
| ≥60 | 131 | 31.1 |
| Mean ± SD. | 49.96 ± 17.03 |  |
| Male | 138 | 32.7 |
| Female | 284 | 67.3 |
| **Ethnicity** |  |  |
| Dalit | 35 | 8.3 |
| Disadvantaged Janajati | 174 | 41.2 |
| Relatively advantaged Janajati | 83 | 19.7 |
| Upper caste group | 130 | 30.8 |
| **Marital Status** |  |  |
| Never married | 22 | 5.2 |
| Currently married | 374 | 88.6 |
| Others* | 26 | 6.2 |
| **Education** |  |  |
| Illiterate | 66 | 15.6 |
| Non-formal schooling | 193 | 45.7 |
| Primary Level (1-5) | 67 | 15.9 |
| Secondary Level (6-10) | 58 | 13.7 |
| HS level and above (13 and above) | 38 | 9.1 |
| **Occupation** |  |  |
| Agriculture | 246 | 58.3 |
| Housework | 118 | 28 |
| Government service | 23 | 5.5 |
| Others** | 35 | 8.3 |

\* Separated, divorced, and widowed
\*\* Private business, foreign employment, driver, private job, and students

### Prevalence of undiagnosed hypertension

The prevalence of undiagnosed hypertension was 30.80% (95% CI: 26.4-35.5). The study participants had mean SBP and mean DBP values of 119.06 ± 19.73 mmHg and 79.11± 11.86 mmHg respectively.

### Behavioral characteristics

More than half of the participants were non-smokers (53.8%), and most had never used smokeless tobacco (81.8%). In terms of alcohol use, a majority reported that they had never consumed alcohol (59.2%). Most participants ate fewer than the recommended servings of fruits and vegetables (79.1%), and almost all were non-vegetarian (98.1%). Over half said they usually added extra salt to their meals (51.2%), and many reported eating processed foods high in salt (69.7%). Nearly half consumed more than 5 grams of salt per day (48.1%), and most did not practice any salt-reduction measures. Refined vegetable oil was the main type of cooking oil used by participants (85.8%). Regarding physical activity, most had a high activity level (91.9%), exceeding 3,000 MET-minutes per week, and very few reported a sedentary lifestyle (98.6%).

**Table 2:** Behavioral characteristics of study participants (n=422)

| **Behavioral characteristics** | **Frequency** | **Percentage (%)** |
| --- | --- | --- |
| **Smoking** |  |  |
| Never smoker | 227 | 53.8 |
| Current smoker | 137 | 32.5 |
| Past Smoker | 58 | 13.7 |
| **Smokeless tobacco** |  |  |
| Never user | 345 | 81.8 |
| Current user | 68 | 16.1 |
| Past user | 9 | 2.1 |
| **Alcohol Consumption** |  |  |
| Never user | 250 | 59.2 |
| Current user | 165 | 39.1 |
| Past user | 7 | 1.7 |
| **Fruits and Vegetables Intake** |  |  |
| Inadequate servings (< 5 Servings) | 334 | 79.1 |
| Adequate Servings (≥ 5 Servings) | 88 | 20.9 |
| **Food habit** |  |  |
| Vegetarian | 8 | 1.9 |
| Non-Vegetarian | 414 | 98.1 |
| **Processed food Consumption** |  |  |
| Yes | 294 | 69.7 |
| No | 128 | 30.3 |
| **Add Table Salt** |  |  |
| Yes | 216 | 51.2 |

| Behavioral characteristics | Frequency | Percentage (%) |
| --- | --- | --- |
| No | 206 | 48.8 |
| **Salt Intake** |  |  |
| ≤ 5 grams | 201 | 47.6 |
| > 5 grams | 203 | 48.1 |
| Don't Know | 18 | 4.3 |
| **Salt Control Measures** |  |  |
| Avoid/minimize achar or papad | 38 | 24.8 |
| Buy low salt alternatives | 3 | 2 |
| Use spice other than salt | 18 | 11.8 |
| Avoid eating outside | 34 | 22.2 |
| No added extra salt from table | 33 | 21.6 |
| Cook meals without adding salt | 27 | 17.6 |
| **Oil consumption** |  |  |
| Mustard oil | 47 | 11.1 |
| Refined vegetable oil | 362 | 85.8 |
| Others* | 13 | 3.1 |
| **Level of physical activity** |  |  |
| Low (< 600 MET) | 11 | 2.6 |
| Moderate (600-2999) | 23 | 5.5 |
| High (>3000MET) | 388 | 91.9 |
| **Sedentary life** |  |  |
| Yes (> 5 hours) | 6 | 1.4 |
| No (≤ 5 hours) | 416 | 98.6 |

\*Vegetable ghee, and animal ghee

### Clinical characteristics, Anthropometric measurement, healthcare seeking behavior of the study participants

Majority of the study participants had diabetes mellitus (30%) followed by respiratory disease (22%) and most of them did not have a family history of hypertension (76.1%). Majority of the study participants (63.7%) had abnormal BMI (Underweight plus overweight plus obesity).

Majority of the participants (96.4%) had their blood pressure checked ever. But more than half (55.6%) did not do regular blood pressure checkup. The reasons for not doing regular blood pressure checkups were no symptoms (70.8%) followed by don’t know (18.6%).

**Table 3:** Clinical characteristics, anthropometric measurement and healthcare seeking behaviour (n=422)

| Variables | Frequency | Percentage (%) |
| --- | --- | --- |
| **Comorbidities (n=49)** |  |  |
| Respiratory Disease | 11 | 22 |
| Diabetes Mellitus | 15 | 30 |
| Others* | 24 | 48 |
| **Family history of Hypertension** |  |  |
| Present | 101 | 23.9 |
| Absent | 321 | 76.1 |
| **Body Mass Index (BMI)** |  |  |
| Underweight (<18.5) | 37 | 8.8 |
| Normal Weight (18.5-22.9) | 153 | 36.3 |
| Overweight (23-24.9) | 82 | 19.4 |
| Obesity ($\geq 25$) | 150 | 35.5 |
| **Ever Measured Blood Pressure** |  |  |
| Yes | 407 | 96.4 |
| No | 15 | 3.6 |
| **Regular Blood Pressure Measure** |  |  |
| Yes | 181 | 44.4 |
| No | 226 | 55.6 |
| **Reasons for not measuring blood pressure regularly (n=226)** |  |  |
| No symptoms | 160 | 70.8 |
| Don't Know | 42 | 18.6 |
| Health center is far | 12 | 5.3 |
| Others** | 12 | 5.3 |

\*Cardiovascular disease, Genitourinary, Mental illness, Thyroid, and Gastro-intestinal (GI) related disease \*\*Interested, and due to fear

### Fhactors associated with undiagnosed hypertension

Age, sex, occupation, current smokeless tobacco, current alcohol consumption, salt intake, level of physical activity, family history of hypertension and regular blood pressure measurement were associated with undiagnosed hypertension at p-value <0.05. Male [AOR=2.2, 95% CI: 1.1-4.2], Salt consumption of more than 5 grams a day [AOR=3.1, 95% CI: 1.8-5.4], and blood pressure not measured regularly [AOR=5.5, 95% CI: 3.1-9.8] remained significant in the multivariate logistic regression model.

**Table 4:**
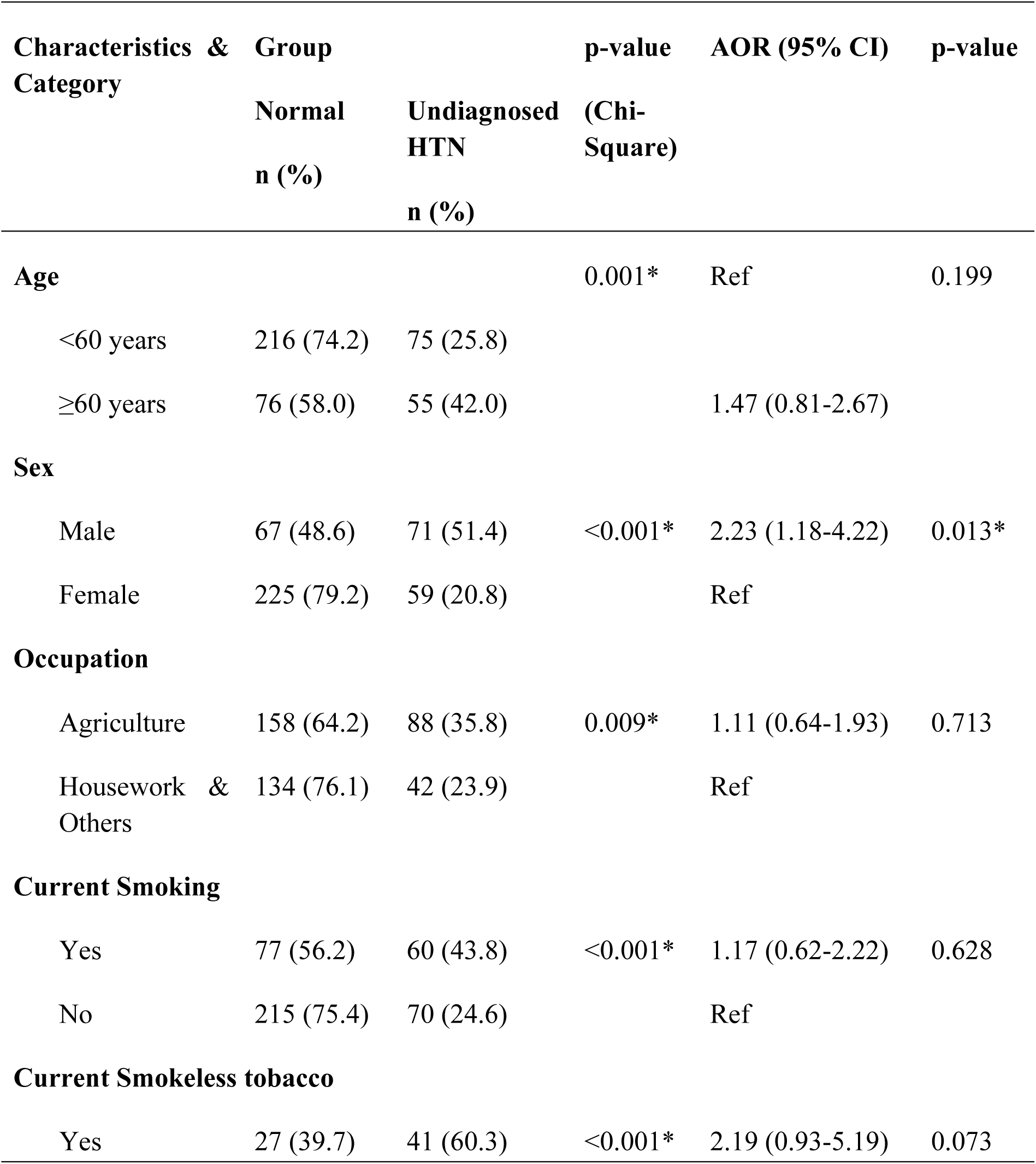
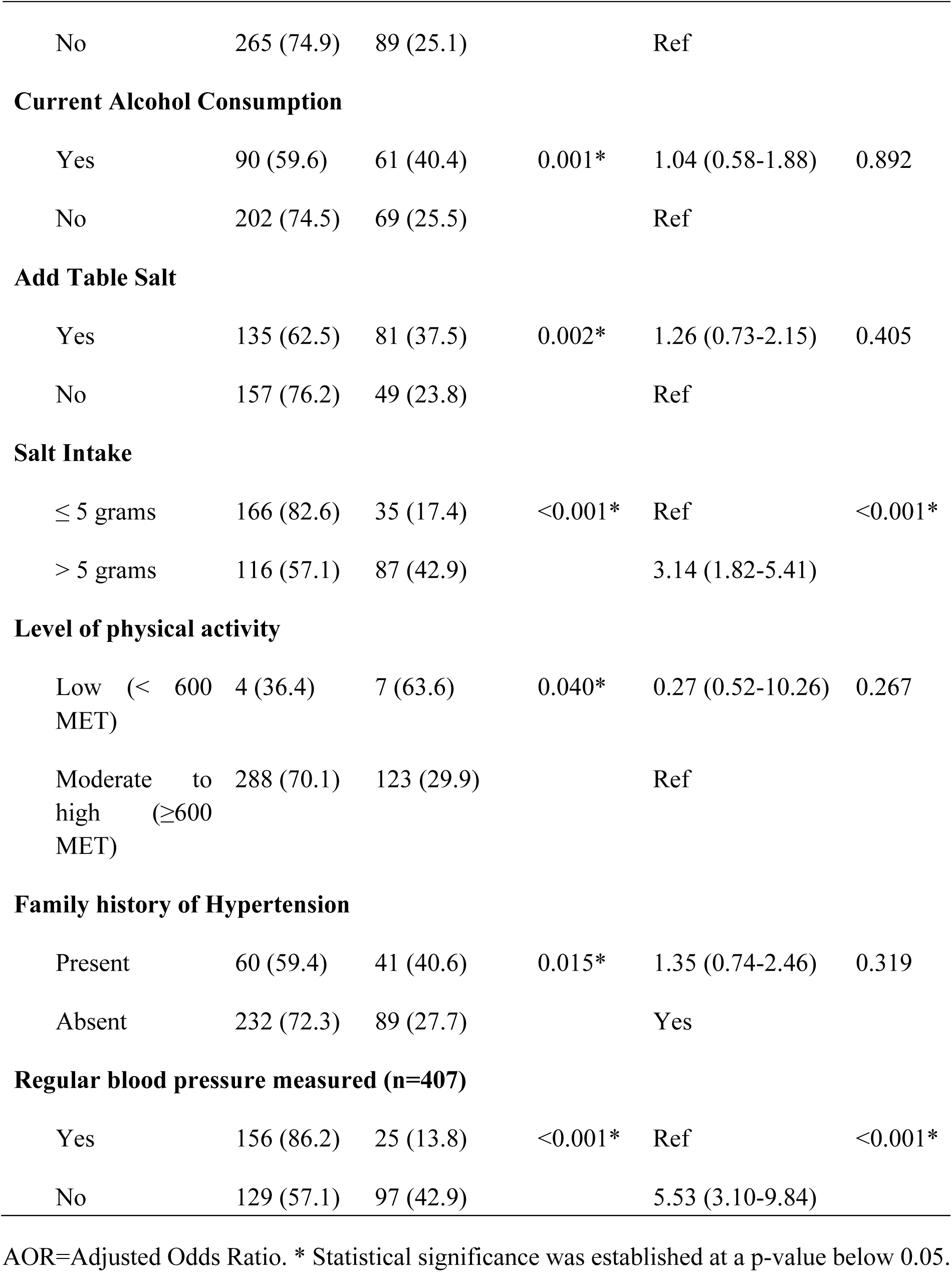
Factors associated with undiagnosed hypertension in bi variate and multivariate logistic regression (n=422)

| Characteristics & Category | Group |  | p-value (Chi-Square) | AOR (95% CI) | p-value |
| --- | --- | --- | --- | --- | --- |
|  | Normal<br>n (%) | Undiagnosed HTN<br>n (%) |  |  |  |
| **Age** |  |  | 0.001* | Ref | 0.199 |
| <60 years | 216 (74.2) | 75 (25.8) |  |  |  |
| ≥60 years | 76 (58.0) | 55 (42.0) |  | 1.47 (0.81-2.67) |  |
| **Sex** |  |  |  |  |  |
| Male | 67 (48.6) | 71 (51.4) | <0.001* | 2.23 (1.18-4.22) | 0.013* |
| Female | 225 (79.2) | 59 (20.8) |  | Ref |  |
| **Occupation** |  |  |  |  |  |
| Agriculture | 158 (64.2) | 88 (35.8) | 0.009* | 1.11 (0.64-1.93) | 0.713 |
| Housework & Others | 134 (76.1) | 42 (23.9) |  | Ref |  |
| **Current Smoking** |  |  |  |  |  |
| Yes | 77 (56.2) | 60 (43.8) | <0.001* | 1.17 (0.62-2.22) | 0.628 |
| No | 215 (75.4) | 70 (24.6) |  | Ref |  |
| **Current Smokeless tobacco** |  |  |  |  |  |
| Yes | 27 (39.7) | 41 (60.3) | <0.001* | 2.19 (0.93-5.19) | 0.073 |
| No | 265 (74.9) | 89 (25.1) |  | Ref |  |
| **Current Alcohol Consumption** |  |  |  |  |  |
| Yes | 90 (59.6) | 61 (40.4) | 0.001* | 1.04 (0.58-1.88) | 0.892 |
| No | 202 (74.5) | 69 (25.5) |  | Ref |  |
| **Add Table Salt** |  |  |  |  |  |
| Yes | 135 (62.5) | 81 (37.5) | 0.002* | 1.26 (0.73-2.15) | 0.405 |
| No | 157 (76.2) | 49 (23.8) |  | Ref |  |
| **Salt Intake** |  |  |  |  |  |
| ≤ 5 grams | 166 (82.6) | 35 (17.4) | <0.001* | Ref | <0.001* |
| > 5 grams | 116 (57.1) | 87 (42.9) |  | 3.14 (1.82-5.41) |  |
| **Level of physical activity** |  |  |  |  |  |
| Low (< 600 MET) | 4 (36.4) | 7 (63.6) | 0.040* | 0.27 (0.52-10.26) | 0.267 |
| Moderate to high (≥600 MET) | 288 (70.1) | 123 (29.9) |  | Ref |  |
| **Family history of Hypertension** |  |  |  |  |  |
| Present | 60 (59.4) | 41 (40.6) | 0.015* | 1.35 (0.74-2.46) | 0.319 |
| Absent | 232 (72.3) | 89 (27.7) |  | Yes |  |
| **Regular blood pressure measured (n=407)** |  |  |  |  |  |
| Yes | 156 (86.2) | 25 (13.8) | <0.001* | Ref | <0.001* |
| No | 129 (57.1) | 97 (42.9) |  | 5.53 (3.10-9.84) |  |

AOR=Adjusted Odds Ratio. \* Statistical significance was established at a p-value below 0.05.

## Discussion

This study revealed that more than a quarter (30.80%) were remained with undiagnosed hypertension. The main associated factors with undiagnosed hypertension were male, salt intake of more than 5 grams a day and not measuring blood pressure regularly.

The prevalence of undiagnosed hypertension in this study was alarming which is similar (30.5%)with other study conducted in in Tertiary referral hospital in Jordan ^24^, Western India 26% ^25^, Northwest Ethiopia 24.8% ^20^. This study finding was lower compared to prior studies in Nepal, one of which utilized nationally representative data and reported a prevalence of undiagnosed hypertension at 50.4% ^11^, and another which utilized DHS 2016 data reported prevalence of 56.9% ^26^. In another study carried out in Nepal, it was found that 69.9% individuals were unaware of their condition who have hypertension ^27^, while study from West Bengal India mentioned it to be 83.86% ^28^ and from Southern Ethiopia mentioned it to be 45.3% ^29^. These differences may be attributed to variations in the characteristics of study participants, sociodemographic disparities, differences in study settings, and variations in sample sizes.

Our findings indicated a higher prevalence compared to a study conducted in eastern part of Nepal (11%) ^30^, and Rural Bangladesh (11.1%) ^31^. These differences might be due to changes in lifestyle and lower level of awareness and asymptomatic nature of hypertension.

Nepal faces high rates of undiagnosed hypertension, marked by significant disparities across demographics and regions. Increasing awareness, regular screenings at basic healthcare centers, and implementing a social health insurance policy may help in curbing this issue ^11^.

This study revealed that male were 2.2 times more prone to undiagnosed hypertension compared to female. The result was consistent with the study conducted in urban area of eastern part of Nepal ^30^, and was also supported by study conducted in Nepal using nationwide survey data^11^. Men were found to have a 2.2 times higher risk of undiagnosed hypertension compared to women. This finding was also consistent with studies conducted outside Nepal, such as those in West Bengal, India ^28^, the Central African Region ^32^, and Southwest Ethiopia ^21^, which demonstrated that men had a greater likelihood of undiagnosed hypertension than females. The significant difference could be due to behavioural factors such as men were more exposed in smoking, alcohol and other risky behavioural activities.

The findings was contradict with the findings from study conducted in India ^33^, which stated that women were at higher risk of being undiagnosed.

This study also unveiled an association between undiagnosed hypertension and daily salt consumption exceeding 5 grams. Similar findings have been reported in studies from eastern Nepal (31), rural Kathmandu (14), as well as international studies from Iran (35) and Malaysia (36). However, accurate assessment of salt intake is challenging, as it relies heavily on participants’ recall and honesty in reporting. Excessive salt consumption contributes to several physiological changes, including water retention, increased systemic peripheral resistance, impaired endothelial function, and alterations in the structure and function of large elastic arteries; all of which collectively elevate blood pressure levels (37).

The likelihood of undiagnosed hypertension increased by 5.5 times among individuals who did not regularly check their blood pressure compared to those who did. This finding was consistent with a study conducted in Tonga (33.3% vs. 19.1%) ^34^. It also corresponds with the guidance provided by the ISH, which suggests that all adults should know their level of blood pressure as the first step in managing hypertension ^35^. Responding to this recommendation, an international hypertension screening program was launched between 2017 and 2019, revealed that 32% of individuals had never undergone blood pressure checkup ^36^. Moreover, studies in China ^37^ and France ^38^ demonstrated that individuals who regularly checked their blood pressure had a higher likelihood of identifying undiagnosed hypertension. Another documented reason is that hypertension often presents as a silent disease with asymptomatic characteristics, leading individuals to forego regular healthcare facility visits ^39^.

There is an evidence from other research ^40, 30, 31, 41^ that some factors including age, smoking, alcohol consumption, low physical activity, having comorbidities and obesity were associated with undiagnosed hypertension but our study showed no association between age, smoking, alcohol consumption, low physical activity, comorbidity, obesity and undiagnosed hypertension. This might be due to variations in study population, study design, operational definition of undiagnosed hypertension, sociocultural variation, geographical variation etc.

## Limitations

The study is cross-sectional, which would limit the ability to establish causality or assess temporal relationships between variables.

## Conclusion

This study found that nearly one-third (30.8%) of participants had undiagnosed hypertension underscoring the urgent need for comprehensive and effective screening strategies to detect and manage hypertension in the broader population of Nepal. Key associated factors included male sex, daily salt intake exceeding 5 grams, and irregular blood pressure monitoring. These findings suggest that a considerable proportion of individuals may be living with hypertension unknowingly. Importantly, undiagnosed hypertension was observed across all age groups and genders, highlighting its widespread risk throughout the population rather than within specific subgroups.

## Data Availability

All data produced in the present study are available upon reasonable request to the authors

## Acknowledgements

We would like to acknowledge the Jwalamukhi rural municipality health section for administrative support and to all the study participants of the study.

## Authors’ contributions

AS, BS, BRK, KR, BB contributed to the conceptualization of the study, study design, data analysis, and manuscript development. BS, BRK, PK & RR contributed to the critical revision of the manuscript. KR and BB supervised and provided mentorship. All the authors contributed to the article and approved the submitted version.

## Conflict of interest

The authors declare that they have no competing interests

### Abbreviations

BMI: Body Mass Index
CVD: Cardiovascular disease
DALY: Disability Adjusted Life Years
DBP: Diastolic Blood Pressure
HICs: High Income Countries
HTN: Hypertension
IRC: Institutional Review Committee
ISH: International Society of Hypertension
LMIC: Low and Middle Income Countries
MET: Metabolic Equivalent of Task
NCD: Non communicable disease
NDHS: Nepal Demographic and Health Services
SAARC: South Asian Association for Regional Cooperation
SBP: Systolic Blood Pressure
SDG: Sustainable Development Goal
SPSS: Statistical Package for Social Science
WHO: World Health Organization

